# A systematic review of prediction accuracy as an evaluation measure for determining machine learning model performance in healthcare systems

**DOI:** 10.1101/2023.06.01.23290837

**Authors:** Michael Owusu-Adjei, James Ben Hayfron-Acquah, Twum Frimpong, Gaddafi Abdul-Salaam

## Abstract

**Background:** Focus on predictive algorithm and its performance evaluation is extensively covered in most research studies. Best predictive models offer Optimum prediction solutions in the form of prediction accuracy scores, precision, recall etc. Prediction accuracy score from performance evaluation have been used as a determining factor for appropriate model recommendations use. It is one of the most widely used metric for identifying optimal prediction solutions irrespective of context or nature of dataset, size and output class distributions between the minority and majority variables. The key research question however is the impact of using prediction accuracy as compared to balanced accuracy in the determination of model performance in healthcare and other real-world application systems. Answering this question requires an appraisal of current state of knowledge in both prediction accuracy and balanced accuracy use in real-world applications including a search for related works that highlight appropriate machine learning methodologies and techniques.

**Materials and methods:** A systematic review of related research works through an adopted search strategy protocol for relevant literature with a focus on the following characteristics; current state of knowledge with respect to ML techniques, applications and evaluations, research works with prediction accuracy score as an evaluation metric, research works in real-world context with appropriate methodologies. Excluded from this review search is defining specific search timelines and the motivation for not specifying search period was to include as many important works as possible irrespective of its date of publication. Of particular interest was related works on healthcare systems and other real-world applications (spam detections, fraud predictions, risk predictions etc).

**Results:** Observations from the related literature used indicate extensive use of machine learning techniques in real-world applications. Predominantly used machine learning techniques were Random forest, Support vector machine, Logistic regression, K-Nearest Neighbor, Decision trees, Gradient boosting classifier and some few ensemble techniques. The use of evaluation performance metrics such as precision, recall, f1-score, prediction accuracy and in some few instances; predicted positive and predicted negative values as justification for best model recommendation is also noticed. Of interest is the use of prediction accuracy as a predominant metric for assessing model performance among all the related literature works indentified.

**Conclusions:** In the light of challenges identified with the use of prediction accuracy as a performance measure for best model predictions, we propose a novel evaluation approach for predictive modeling use within healthcare systems context called PMEA (Proposed Model Evaluation Approach) which can be generalized in similar contexts. PMEA, addresses challenges for the use of prediction accuracy with balanced accuracy score derived from two most important evaluation metrics (True positive rates and True negative rates: TPR, TNR) to estimate more accurately best model performance in context. Identifying an appropriate evaluation metric for performance assessment will ensure a true determination of best performing prediction model for recommendation.

## INTRODUCTION

A key component in disease treatment is estimating outcome after treatment is initiated. An outcome is driven mainly by two critical issues; patient compliance and efficient treatment strategies on the part of healthcare givers. Developing effective and efficient strategies [1] for managing severely ill patients remains a major challenge for healthcare providers. Associated morbidity and mortality as undesirable consequence of undetected and insufficient care management practices of uncontrolled blood pressure by individuals is an important justification for the adoption of predictive learning technique use capable of identifying important correlated factors associated with the incidence of hypertension and thereby assist in providing real-time solutions to low detection rates among many segments of society. Increasing data generation capacities coupled with availability of tools for data collection arising from increasing use of automated systems and Internet of things (IoT) as an emerging paradigm [2], [3] involving human interactions and interconnection of devices has contributed to the availability of large volumes of datasets being witnessed today. Characteristically, healthcare systems are associated with generation of large volumes of datasets brought about as a result of connected medical device use such as remote patient monitoring and virtual assistant devices for use in areas such as blood pressure, pulse, heart rate, diabetic monitors etc. Others include, connected contact lenses, glucose monitors, wearables, fitness tracking devices, virtual healthcare assistants, virtual dispensing assistants etc. Generated data from many of these applications have been explored and exploited in many research works to generate patterns of change using various Predictive machine learning (ML) approaches also referred to as non-clinical approaches [4] to enhance disease diagnosis and treatment outcome. The performance of these predictive models have become subject for many research works throughout history after evaluation.

Evaluation in general involves three important qualities which are systematic, assessment and the determination of value, worth and significance. Systematic connotes an interpretation which is structured to give meaning. Different predictive techniques involves the use of different or same evaluation metrics [5]. Example, predictive evaluation metrics for ML techniques in Classification analysis may be the same or differ from those used in Regression analysis depending on the problem under consideration. The challenge here is when to use what and for what reason and to what benefit. Identifying the appropriate domain for its use and for what reason such as evaluate performance for optimization or estimating the number of correctly classified patients for treatment default, number of patients with certain types of diseases etc is a better use of predictive models. In this review, we offer a thorough discussion on various performance evaluation metrics in line with the key research question: Effects of using prediction accuracy score as compared to balanced accuracy in the context of identifying best machine learning model for predictive performance.

### Review Contribution

This paper highlights an important ingredient in the choice of best machine learning model for prediction and places this choice under context. We also make an assertion that the supposedly higher prediction accuracy scores as obtained in some research findings when compared with balanced accuracy scores of studies using similar ML techniques in the same context creates an erroneous impression of high performing models among individual ML techniques and for this reason the choice of best performing ML model based on prediction accuracy is problematic if context and purpose for prediction modeling is not considered. We have used only one evaluation metric (accuracy score) many others remain, we therefore encourage further discussions on the appropriate use of all the other evaluation metrics for emphasis.

### Review Methodology

Our approach was to adopt guidelines emphasized in the preferred reporting items for systematic reviews and meta-analyses (PRISMA) protocol. These protocols were; designing the research question, adopting searches and search strategy, developing inclusion and exclusion criteria, designing data extraction plan to synthesis and draw conclusions, quality assessment criteria rule and developing strategies to analyzed the collected data.

### Search Strategy

Literature used were obtained from the following sources; PuMed, Google scholar, Web of science indexed journals, Scopus indexed journals (Springer nature, Hindawi, Elsevier, ScienceDirect, IEEEAccess, IEEEXplore) and many others. Search words included; predictive modeling in healthcare systems, machine learning prediction accuracy score, disease diagnosis with machine learning, machine learning prediction of diseases (chronic kidney, hypertension, breast cancer, machine learning model performance evaluations, fraud detection with machine learning, detection of spam messages with machine learning, machine learning prediction with balanced accuracy score, dealing with class imbalance in machine learning etc. Our search period was started from 2016 to ensure access to most materials since ML use in healthcare has been limited since its inception.

### Inclusion Criteria

Our inclusion criteria for relevant articles were; model performance evaluation metrics, evaluation with accuracy scores, prediction with ML methods (techniques), ML applications in healthcare, ML use in healthcare (diagnosis, treatments, disease management), fraud detections, spam detections, risk predictions, junk mail predictions, ML in disease treatment default, deep learning applications in healthcare and many others.

### Exclusion Criteria

Excluded from the search criteria were; ML application articles without performance evaluation, articles considered to be outside the realm of real-world application, articles with duplicate findings, articles with findings inconsistent with stated research objectives and reviewed articles.

### Data Extraction Plan

To assist in extracting relevant information from the sourced documents, every single article downloaded were placed in Mendeley Desktop including source documents from non-academic websites (industrial webpages with relevant information).

### Quality Assessment

Our quality assessment procedure was to follow through with all protocols stated above and this resulted in the use of 68.6% of total articles sourced (meeting all inclusion and exclusion criteria) as described in figure3 and figure4.

**Figure1.**
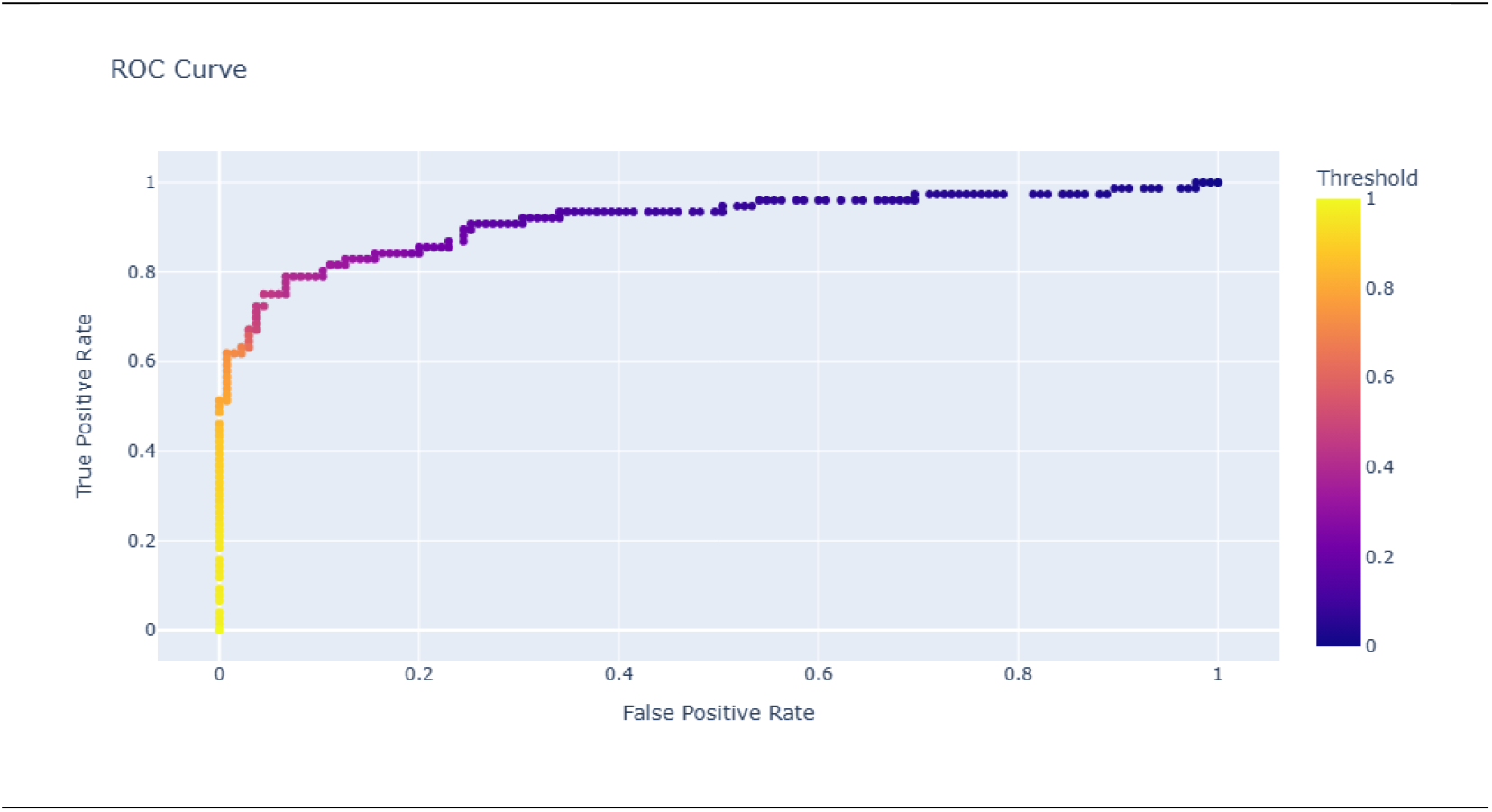
roc_auc curve showing Threshold levels.

**Figure2.**
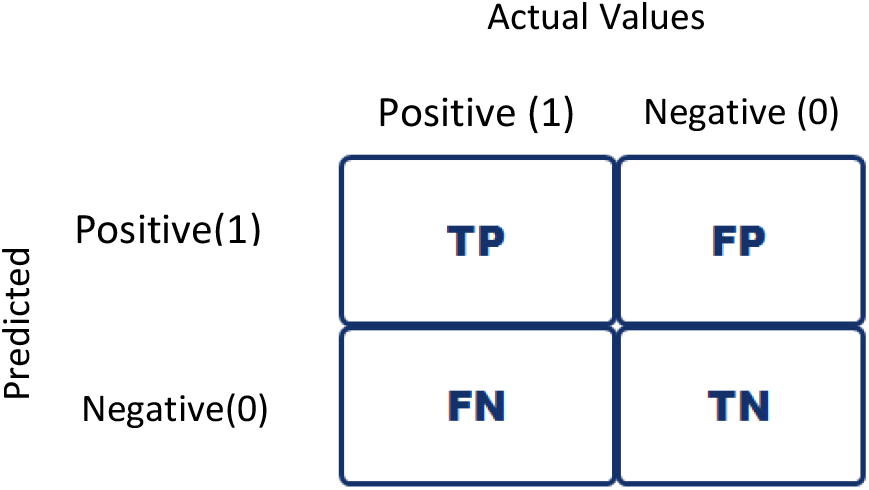
Confusion matrix

**Figure3.**
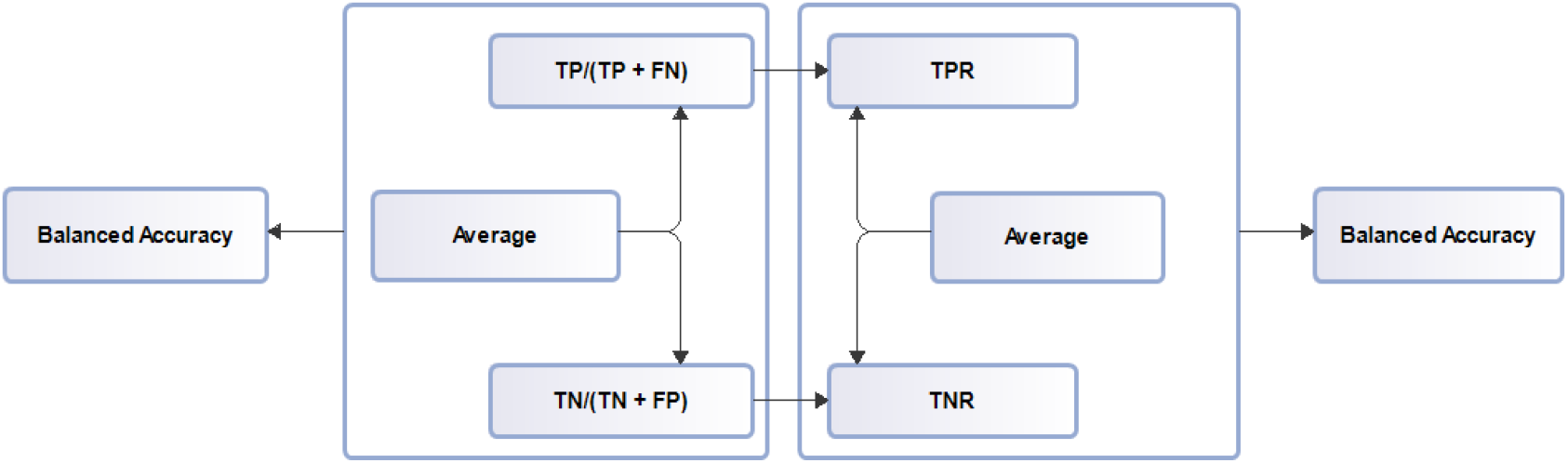
Balanced accuracy score

**Figure4.**
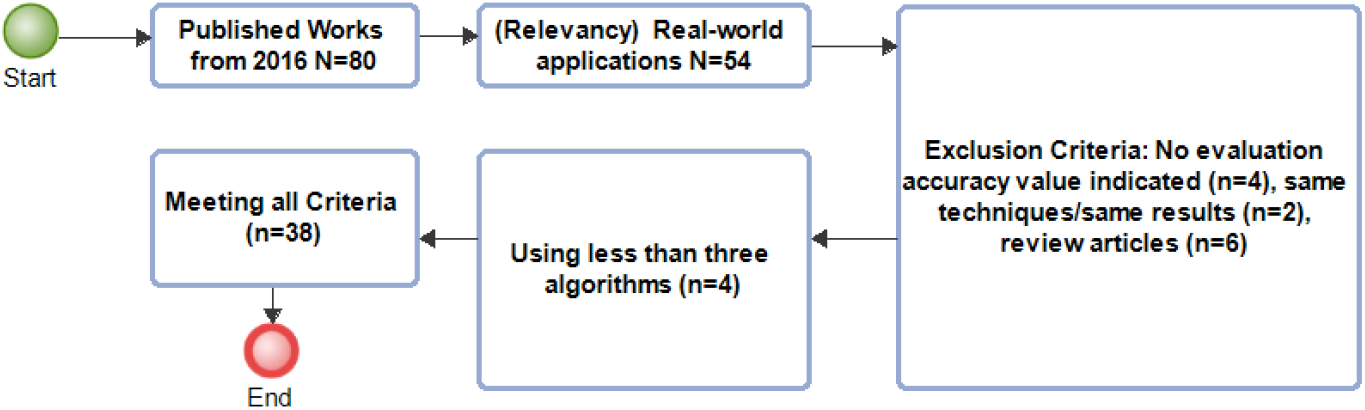
Flowchart diagram detailing selection and exclusion criteria.

### Evaluation metrics in Classification

Brief description of performance evaluation metrics used in most machine learning applications for classification to demonstrate metric use and reasons for its use.

### Prediction Accuracy

In ML, prediction accuracy defines how well a model performs at predictions on unseen data. Prediction accuracy is only a fraction of model predictions that are correct [6]. Prediction accuracy is illustrated as

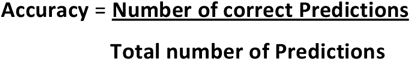

Subsequently in classification, accuracy is calculated in terms of positive and negative predictions.

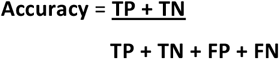

Where

TP = True positives, TN= True negatives, FP = False positives, FN = False negatives

### Receiver operating characteristic curve (roc_auc)

Measures the performance of ML model’s ability to differentiate between classes. A higher roc_auc curve score closer to 1 indicates a favourable model performance at predicting 0 as 0 and 1 as 1. Some of the terms used in roc_auc curve are TPR (True positive rates/ Recall/Sensitivity)

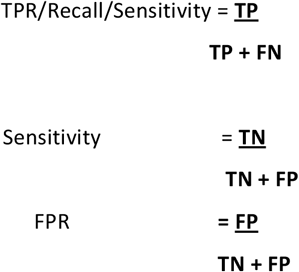

Where FPR = False positive rates

### ROC Curve

Decrease in threshold leads to increase in more positive values and an increase in sensitivity with a decrease in specificity. Conversely an increase in threshold leads to more negative values and a high specificity with low sensitivity [7].

### Confusion matrix

Classification performance metric which consists of combination of predicted and actual values presented in a table format. Confusion matrix is the foundation on which precision, recall, roc_auc, specificity and prediction accuracy is derived.

### Log-loss

Measures the closeness of the prediction probability to the corresponding actual value or true value (0 or 1). A higher log-loss is indicative of divergence of the prediction probability from the actual value.

### Precision

Refers to the model’s identification of relevant data points. Its ability to identify true data points that are positive and classified by the model also as positive. False negative predictions are data points the model identifies as negative but are truly positive (false alarm).

### Recall

A models ability to identify all relevant class instances in a dataset. In certain situations, precision and recall can be combined to achieve optimal solution to a problem such as identifying all patients labelled as defaulters to disease treatment. This will lead to a high recall value but a low precision score.

### F1-score

The harmonic mean of precision and recall to achieve optimal solution (combining precision and recall).

### Evaluation metrics in Regression

Some of the evaluation metrics used in regression analysis are as follows;

- Mean Squared Error (MSE)
- Mean Absolute Error (MAE)
- Mean Absolute Percentage Error (MAPE)
- Root Mean Squared Error (RMSE)
- Root Mean Error (RME)
- Adjusted R-Squared (Adjusted R^2^)

### Balanced Accuracy

A metric used in imbalanced datasets for evaluation performance. It is the average of sensitivity and specificity.

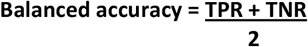

Where

TPR =True Positive Rates

TNR = True Negative Rates

### Balanced accuracy process diagram

The concept of ML application in healthcare practice continue to receive attention both from healthcare providers and policy formulators. The challenge however is its implementation due largely to lack of understanding and confidence on the part of healthcare users in ML techniques. It is important that the development and evaluation of used ML techniques are made transparent and interpretable to allay any doubt about its usability in healthcare systems. Predictive model evaluation especially in healthcare and other real-world application systems must take into account the peculiarity of its dataset especially when assessing predictive model performance [8]. General prediction accuracy score show results obtained from both observed and predicted values. It is predominantly used in classification problems where there are no dataset imbalances and no skewed dataset examples. However, one of the challenges identified in related research works is its use as a performance metric to estimate best machine learning model technique in real world applications such as healthcare systems where dataset skewedness and class imbalance is prevalent. The challenge of using prediction accuracy as a measure of model performance is mentioned in a related review work that examined the prospects of machine learning use in clinical outcomes [9]. This concern regarding prediction accuracy score use is also shared in a study of disease diagnosis with 20 machine learning techniques such as Naïve Bayes, Support Vector Machine (SVM), K-Nearest Neighbors (KNN), perceptron, Light Gradient Boosting Machine, extreme Gradient Boosting etc and to address this challenge, the evaluation metric of f1-score was applied [10]. Prediction accuracy scores obtained ranged between 0.49-0.77 for the various predictive techniques but the f1-score ranged between 0.47-0.82. In their systematic study of prediction accuracy for type2 Diabetes mellitus, a comparative analysis of a related work with the use of prediction accuracy is copiously illustrated for emphasis [11]. A review study of artificial intelligence in disease diagnosis mentioned prediction accuracy as one of the evaluated parameters in related studies of interest [12]. Similarly, a comparative review study of disease prediction with supervised ML techniques also identified various related studies with prediction score as performance metric [13]. Similar use for prediction accuracy [14] in assessing best ML technique for breast cancer prediction recorded an accuracy score of 98.7% for techniques such as decision trees and other ensemble techniques. ML principles and applications in real world systems have also been explored [15]. An automatic prediction system for diabetic patients using dataset of females and several ML techniques for an explainable artificial intelligence [16] concluded with prediction accuracy score of 81% and an auc score of 0.84. Furthermore studies such as prediction of an occurrence of pressure ulcer nursing adverse event [17] using four different ML techniques namely; Decision trees, Support Vector Machines, Random Forest and Artificial Neural Networks achieved a prediction accuracy scores of 94.94% for Support vector machine, 97.93% for Decision trees, 99.88% for Random Forests and 79.02% for Artificial Neural Networks. Determination of best ML algorithms for identifying mental health problems [18] in its early stage with ML techniques such as Logistic Regression, Gradient Boosting, Neural Networks, K-Nearest Neighbor, and Support Vector Machine, as well as an ensemble techniques showed an overall prediction accuracy score of 88.80% which was achieved by Gradient Boosting. Additional studies to predict heart disease with ML algorithms such as K-Nearest Neighbors (KNN), Naive

Bayes and Random Forest singled out Random Forest as the best performing classifier with prediction accuracy score of 95.63% [19]. A review of approaches and trends [20] regarding the use of graph machine learning technique in disease prediction, made preference for the adoption of graph-based neural network technique as recommended approach because of its potential benefit in medical diagnosis, disease treatments and prognosis of diseases. Further studies for ML use in cardiovascular disease prediction with learning techniques such as support vector machine and convolutional neural networks together with boosting classifiers produced prediction roc_auc scores of range 0.81-0.97 [21]. Diagnosis of breast cancer with learning techniques such as linear discriminant analysis (LDA) and Support vector machine (SVM) for various roles had prediction accuracy readings as 99.2% and 79.5% [22]. However, the prediction of breast cancer with Decision tree and Random forest techniques [23] showed prediction accuracy scores of 91.18% and 95.72% respectively. Additional ML application for decision support [24] in the detection of breast cancer through feature selection with ML techniques as k-Nearest Neighbor, linear discriminant analysis, and probabilistic neural network yielded an accuracy score of 99.17%. Furthermore [25], prediction of breast cancer with ML based framework using ML techniques; Random Forest, Gradient Boosting, Support Vector Machine, Artificial Neural Network, and Multilayer Perception to achieve better classification accuracy using correlation-based feature selection together with recursive feature elimination extraction resulted in prediction accuracy score of 99.12%. Similarly, with weighting feature and backward elimination feature selection approaches [26], application of Random forest ML technique, to create computer-aided diagnostic system that distinguishes breast cancer tumors between malignant and benign yielded prediction accuracy score of 99.7% and 99.82% respectively. Achieving a higher precision and prediction accuracy using K-fold cross-validation with all features in model 2, all features without validation in model 1, with feature selection for model 3 and feature selection together with cross-validation [27] for model 4 using ML techniques; logistic regression, support vector machines, Naive Bayes, Decision trees and k-nearest neighbor, produced different prediction accuracy scores at each stage. Highest accuracy scores of importance recorded were; 98.83% for support vector machine, 97.17% for K-Nearest Neighbor and 97.88% for Logistic regression. Similarly, ML based model for early stage heart disease prediction with ML techniques such as support vector machine, K-nearest neighbor, random forest, Naive Bayes, and decision tree with different feature selection techniques (chi-square, ANOVA, and mutual information) for a determination of best fit model along with appropriate features segmented into three categories concluded that Random forest had the highest prediction accuracy of 94.51% [28].

A related study on choice of best ML model for the prediction of [29] breast cancer through the creation of a web system to facilitate prediction analysis also had prediction accuracy scores of 0.98 for Artificial Neural Network, 0.98 for Decision tree classifier, 0.99 for K-Nearest Neighbor, 0.98 for Logistic regression and 1.0 for Support vector machine. Furthermore risk prediction and diagnosis [30] of breast cancer through a comparative analysis of ML techniques to assess model efficiency and effectiveness with respect to prediction accuracy, precision, sensitivity and specificity proved that support vector machine had the highest prediction accuracy performance of 97.13% with the least error rate. Related study [31] to predict and diagnose breast cancer using ML techniques for the determination of best model with respect to evaluation metrics such as confusion matrix, accuracy and precision proved that Support Vector

Machine among other ML techniques (Random Forest, Logistic Regression, Decision tree (C4.5) and K-Nearest Neighbors) achieved the greatest prediction accuracy score of 97.2%. The continuous use of models such as Support vector machines, Logistic regression and Random forest and Clustering in classification problems such as chronic disease diagnosis is emphasized in a related study that found them to be useful [32]. Similarly, the prediction of treatment trend for patients suffering from hypothyroidism using sodium levothyroxine with ML techniques showed that using extra-trees achieves better prediction accuracy of 84%. [33]. Following from this [34] is a predictive study of chronic kidney disease prediction with three ML techniques namely; Random forest, Support Vector machine and Decision tree together with recursive feature elimination technique. This study showed different prediction accuracy scores in situations where feature selection is used and others where feature selection is not used. Prediction accuracies recorded with feature selection techniques are as follows; 99.8% for Random forest, 95.5% for Support vector machine and 98.6% for Decision tree. Additional studies on predictive modeling of chronic diseases such as sclerosis progression and outcomes using ML techniques [35] using ML techniques such as K-nearest neighbor, Support vector machine, Decision tree and Logistic regression concluded with performance evaluation metrics such as area under the curve score (auc), sensitivity, specificity, geometric mean and f1-score. Furthermore studies [36] for the detection of chronic kidney disease that shows important correlations or predictive attributes using ML techniques (k-nearest neighbors, random forest, and neural networks) and 24 features used accuracy, root mean squared error (rmse) and fi-score measure as evaluation parameters to achieve a predicted accuracy score of 0.993 with Random forest classifier. Additional research to identify advanced chronic kidney diseases with ML techniques; generalized linear model network, random forest, artificial neural network and natural language processing through a combination of different datasets [37] showed improved prediction performance in accuracy scores as reported. Prediction accuracy scores for the used ML techniques were; both for training data and testing data: Logistic regression 81.8% and 81.9%, Random forest 91.3% and 82.1%, Decision tree 86.0% and 82.1%. Its conclusion recommends an improvement on these prediction accuracy scores. The application of deep learning in the prediction and classification of hypertension with blood pressured related variables for which those positive for hypertension was 1883 and those without hypertension were 6266 [38], a comparative performance evaluation between deep neural network and decision tree classifier with four different datasets showed the following prediction accuracies; Deep neural network: (0.75, 0.739, 0.743, 0.743) and for Decision tree: (0.676, 0.684, 0.690, 0.680). Further risk prediction studies aid at improving prediction performance with reliable techniques [39] for cardiovascular diseases using ML techniques such as K-nearest neighbor and Multi-layer perceptron (MLP) showed a prediction accuracy of 82.47% for MLP. Similarly, a related study [40] on the prediction of hypertension using features such as patient demographics, past and current patient health condition and medical records for the determination of risk factors through the use of artificial neural network showed prediction accuracy of 82%. Understanding disease symptoms is one sure way of effectively controlling and managing its treatment outcome. Predictive modeling [41] of heart disease risks and its symptoms using ML techniques will ensure effective patient care. Implementation of heart disease risk prediction using six ML techniques (support vector machine,

Gaussian Naive Bayes, Logistic regression, light gradient boosting model, extreme gradient boosting and Random forest) showed the following predicted accuracies; 80.23%, 78.68%, 80.32%, 77.04%, 73.77% and 88.5% respectively. A population level-based approach [42] for the prediction of hypertension using ML techniques for a study example of 818603 with 82748 constituting 10.11% positive for hypertension. Using the following ML techniques (extreme Gradient Boosting, Gradient Boosting Machine, Logistic Regression, Random forest, Decision tree and Linear Discriminant Analysis), predicted accuracy scores obtained were; 90% for (extreme Gradient Boosting, Gradient Boosting Machine, Logistic Regression and Linear Discriminant Analysis) as compared to 89% for Random forest and 83% for Decision tree.

### Accuracy score in non-health settings

Related research perspectives in other real-world applications such as spam message detection, fraud detection and risk estimation/forecasting are explored in this section. The risk of spam messaging and its impact on business operations are far reaching. Systems have been hacked, ransoms have been paid, important data have also been destroyed by viruses and many more. Applying effective, efficient ML modeling techniques that identifies important characteristics for the detection and subsequent prevention or destruction of threats posed continue to engage research attention. A study to detect spam threats [43] in emails and IoT platforms using Naıve Bayes, decision trees, neural networks and random forest including other techniques, prediction accuracy and precision for Suppost Vector Machine and Naive Bayes recorded were (prediction accuracy for Support vector machine; 96.9%, precision 93.12% and prediction accuracy for Naive Bayes; 99.46%, precision 99.66%) higher than the others indicating better performance in spam detections. Similarly, a transformer-based embedding with ensemble learning techniques for spam detection also showed prediction accuracy score of 99.91% [44]. Furthermore application [45] of a hybrid algorithm for the detection of malicious spam messages in email using ML techniques such as Naive Bayes, Support vector machines, Logistic Regression and Random Forest show predicted accuracy scores of 96.15% for Naive Bayes, 96.15% for support vector machine, 98.08% for Logistic regression and 95.38% for Random forest respectively. Evaluation of performance [46] using deep learning approach for automatic short message service (sms) spam classification with the following ML techniques; Naive Bayes, BayesNet, C4.5, J48, Self-organizing map and Decision tree showed predicted accuracy scores as 89.64%, 91.11%, 80.24%, 79.2%, 88.24% and 75.76% respectively. Comparative evaluation to improve prediction accuracy [47] of two ML models; support vector machine and random forest for the detection of junk mail spam showed prediction accuracies of the two models as; Support vector machine 93.52% and Random forest 91.41%. Related to improving prediction accuracy is the issue of improving training time and reducing prediction error rate. ML based hybrid bagging technique application [48] using random forest and decision tree (J48) for the analysis of email spam detection showed bagging technique achieve 98% prediction accuracy. Other performance metrics evaluated include true negative rates, false positive rate and false negative rate, precision, recall and f-measure (f1-score). Similarly, recent increase in online transactions including online payments has also increased the risk of credit card fraud, ML based credit card fraud detection system [49] using genetic algorithm as feature selection with the following learning techniques (Decision Tree, Random Forest, Logistic Regression, Artificial Neural Network, and Naive Bayes showed that applied genetic algorithm feature selection led to a predictive accuracy score of 100% for both Decision tree and Artificial neural network. Related to study [49] is financial fraud detection system in healthcare using ML techniques including deep learning that addresses the challenge of credit card fraud monitoring [50]. Applying the following ML techniques (Naive Bayes, Logistic Regression, K-Nearest Neighbor, Random Forest, and Sequential Convolutional Neural Network) resulted in the following predicted accuracy scores; 96.1%, 94.8%, 95.89%, 97.58%, and 92.3% respectively. Strategies have been adapted and adopted to deal with the challenge of fraud detection by various organizations. One such solution is provided by [51] which implements ML based self-analyzing system that flags potential fraudulent activities for review. Case study approach [52] for a review of ML techniques (logistic regression, decision tree, random forest, K-Nearest Neighbor and extreme Gradient Boosting) in credit card fraud detection evaluated best model prediction performance using accuracy, recall, precision and f1-score metrics. The study identified Logistic regression and K-nearest Neighbor as best performing classifiers. Implementation of fraud detection tools [53] to identify anomalies on financial applications using outlier detection techniques such as Local outlier factor, Isolation factor and Elliptic envelope and ML techniques (Random forest, Adaptive boosting and extreme gradient boosting) showed predicted accuracy score of 0.9995. An unplanned modeling [54] for medical visits by patients suffering from diabetes with ML techniques; logistic regression, support vector machine, linear discriminant analysis, quadratic discriminant analysis, extreme gradient boosting, neural networks and deep neural network obtained a balanced accuracy score of 65.7%. Similarly, predicting length of stay [55] from admission to a clinical ward with ML techniques such as random forest, decision trees, support vector machine, multi-layer perceptron, adaboost and gradient boost concluded with random forest as the best performing technique with a balanced accuracy score of 0.72 at the initial stage of admission and 0.75 in-admission. However, an up-sampling approach [56] for breast cancer prediction using k-nearest neighbor, decision tree, random forest, neural networks, support vector machine and extreme gradient boosting obtained a balanced accuracy score of 97.47%.

### Related works summary

The systematic review of related research works had key objectives and among them was the search for literature with the following characteristics; a focus on current state of knowledge with respect to ML techniques, applications and evaluations, research works with prediction accuracy score as an evaluation metric, research works in real-world context with important methodologies. Excluded from this review article search is defining specific search timeline and the motivation for not specifying search period was to include as many important works are possible irrespective of its date of publication. Of particular interest was related works on healthcare systems and other real-world applications (spam detections, fraud predictions, risk predictions etc). A summary of identified characteristics among selected reviewed literature with emphasis on prediction accuracy score as performance metric is presented in table1. Literature search sources were; Google scholar and other online journal databases such as IEEE, puhmed, hindawi journals, BioMed central, Pmc, Elsevier, Sciencedirect, organizational websites, online libraries and many other journals. A total of 80 articles were screened for (relevancy), determined inclusion criteria was for related works in healthcare practice that had used predictive machine learning either in disease diagnosis, prediction, risk or treatment assessment. Literature of related works with ML applications in other relevant settings such as in information technology industry (example; spam detection in mails, sms spamming) were also considered. No time frame exclusion criteria was used, but about 80% of selected materials were mainly published works between 2016-2022 and a handful in 2023. Statistical and graphical observations about the reviewed literature are shown in Figure5 and Figure6 which also includes a flowchart diagram of the selection process shown in Figure4. Observations noticed in related literature used indicate extensive use of various ML techniques in real-world applications for various reasons some of which as decision support system. Predominantly used techniques include Random forest, Support vector machine, Logistic regression, K-Nearest Neighbor, Decision trees, Gradient boosting classifier and some few ensemble techniques. The use of evaluation performance metrics such as precision, recall, f1-score, prediction accuracy and in some few instances; predicted positive and predicted negative values. Of interest is the use of prediction accuracy as a predominant metric for assessing model performance found among all the related literature reviewed.

**Table1.** Reviewed literature descriptions

| Reference no | Research type | Methodology | Evaluation metric | Score value |
| --- | --- | --- | --- | --- |
| [8] | Disease diagnosis | Naïve Bayes, Support Vector Machine (SVM), K-Nearest Neighbors (KNN), perceptron, Light Gradient Boosting Machine, extreme Gradient Boosting. | accuracy | 0.49-0.77 |
| [12] | Breast cancer prediction | Decision trees, ensemble techniques | accuracy | 98.7 |
| [14] | Diabetic prediction |  |  | 81 |
| [15] | Pressure ulcer nursing prediction | Decision trees, Support Vector Machines, Random Forest and Artificial Neural Networks | accuracy | 94.94, 97.93, 99.98, 79.02 |
| [16] | Identify mental health | Logistic Regression, Gradient Boosting, Neural Networks, K-Nearest Neighbor, and Support Vector Machine, as well as an ensemble techniques | accuracy | 88.80 |
| [17] | Heart disease prediction | K-Nearest Neighbors (KNN), Naive Bayes and Random Forest singled out Random Forest | accuracy | 95.63 |
| [19] | Cardiovascular disease prediction | support vector machine, convolutional neural networks boosting classifiers | accuracy | 0.81-0.97 |
| [20] | Breast cancer diagnosis | linear discriminant analysis (LDA) and Support vector machine (SVM) | accuracy | 99.2, 79.5 |
| [21] | Breast cancer prediction | Decision tree, Random forest | accuracy | 91.18, 95.72 |
| [22] | Detection of breast cancer | K-Nearest Neighbor, linear discriminant analysis | accuracy | 99.17 |
| [23] | Prediction of breast cancer | Random Forest, Gradient Boosting, Support Vector Machine, Artificial Neural Network, and Multilayer Perception | accuracy | 99.12 |
| [24] | Differentiate between malignant and benign tumors diagnosis | Random forest with weighting and backward elimination feature techniques | accuracy | 99.7, 99.82 |
| [26] | early stage heart disease prediction | Support vector machine, K-nearest neighbor, Random forest, Naive Bayes, and Decision tree | accuracy | 94.51 |
| [27] | Prediction of breast cancer | Artificial Neural Network, Decision tree classifier, K-Nearest Neighbor, 0.98, Logistic regression, Support vector machine | accuracy | 0.98, 0.98, 0.99, 0.98, 1.0 |
| [28] | Breast cancer risk prediction | Support vector machine | accuracy | 97.13 |
| [29] | Breast cancer diagnosis | Support vector machine, Random Forest, Logistic Regression, Decision tree (C4.5) and K-Nearest Neighbors | accuracy | 97.2 |
| [31] | Treatment trend prediction for hypothyroidism | Extra trees | accuracy | 84 |
| [32] | Chronic kidney disease prediction | Random forest, support vector machine, decision tree | accuracy | 99.8, 95.5, 98.6 |
| [33] | Chronic disease progression (sclerosis) | K-nearest neighbor, support vector machine, decision tree, logistic regression | auc |  |
| [34] | Chronic kidney disease detection | K-nearest neighbor, random forest, neural networks | accuracy | 0.993 |
| [35] | Advanced chronic kidney disease prediction | Logistic regression, random forest, decision tree | accuracy | 81.9, 82.1, 82.1 |
| [36] | Prediction of hypertension | Deep neural network, decision tree | accuracy | 0.75, 0.69 |
| [37] | Risk prediction (hypertension) | k-nearest neighbor, multi-layer perceptron | accuracy | 82.47 |
| [38] | Prediction of hypertension | Artificial neural network | accuracy | 82 |
| [39] | Heart disease risk prediction | support vector machine, Gaussian Naive Bayes, Logistic regression, light gradient boosting model, extreme gradient boosting and Random forest | accuracy | 80.23, 78.68, 80.32, 77.04, 73.77, 88.5 |
| [40] | Hypertension prediction | extreme Gradient Boosting, Gradient Boosting Machine, Logistic Regression, Random forest, Decision tree and Linear Discriminant Analysis | accuracy | 83-90% |
| [41] | Spam detection | Naive Bayes, decision tree, neural networks, random forest, support vector machine | accuracy | 96.9-99.66 |
| [42] | Spam detection | ensemble | accuracy | 99.91 |
| [43] | Malicious spam in mails | Naive Bayes, support vector machine, logistic regression and random forest | accuracy | 96.15, 96.15, 98.08, 95.38 |
| [44] | Sms spam classification | Naive Bayes, BayesNet, C4.5, J48, Self-organizing map and Decision tree | accuracy | 89.64, 91.11, 80.24, 79.2, 88.24, 75.76 |
| [45] | Junk email detection | Support vector, random forest | accuracy | 93.52, 91.41 |
| [46] | Email spam detection | Bagging, random forest, decision tree (J48) | accuracy | 98 |
| [47] | Credit card fraud detection | Decision Tree, Random Forest, Logistic Regression, Artificial Neural Network, and Naive Bayes | accuracy | 100 |
| [48] | Financial fraud detection in healthcare | Naive Bayes, Logistic Regression, K-Nearest Neighbor, Random Forest, and Sequential Convolutional Neural Network | accuracy | 96.1, 94.8, 95.89, 97.58, and 92.3 |
| [51] | identify anomalies on financial applications | Random forest, Adaptive boosting and extreme gradient boosting | accuracy | 0.9995 |
| [54] | Unplanned medical visit | Logistic regression, support vector machine, neural network, deep neural network, extreme gradient boosting, linear discriminant analysis, quadratic discriminant analysis | Balance accuracy | 65.7 |
| [55] | Patient length of stay | Random forest, decision tree, support vector machine, multi-layer perceptron, adaboost and gradient boosting | Balanced accuracy | 0.75 |
| [56] | Breast cancer prediction | k-nearest neighbor, random forest, decision tree, neural network, support vector machine and extreme gradient boosting | Balanced accuracy | 97.47 |

**Figure5.**
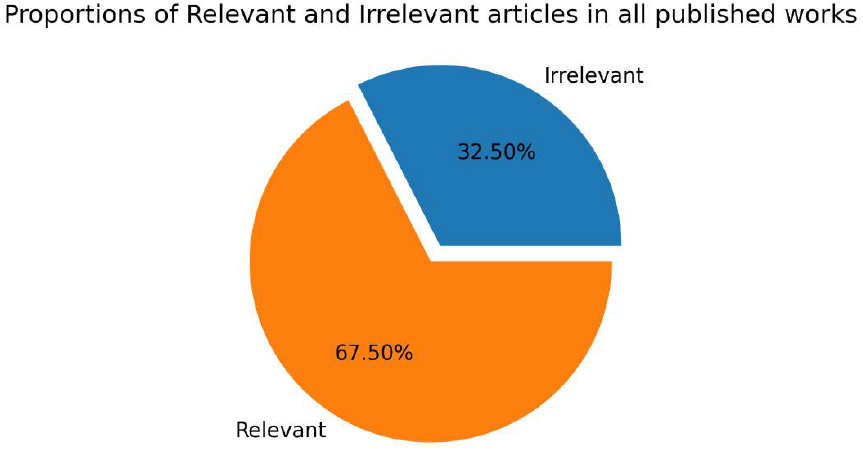
Shared distribution between relevant and irrelevant articles

**Figure6.**
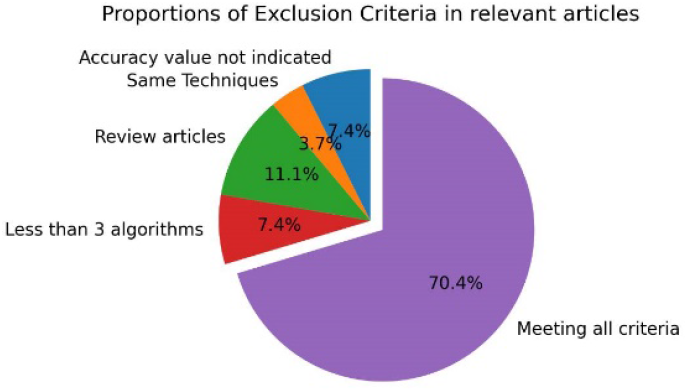
Distribution share among exclusion criteria

### Selection of related works process flowchart

### Strengths and Weaknesses Identified in reviewed literature

In many of the literature reviewed, the pattern of higher prediction accuracy score is identified and there is strong indication of the use of more than one predictive techniques for comparative analysis. The use of predictive modeling in diseases, disease diagnosis, treatment outcome including diseases of public concern whether in e-mail spam predictions, fraud detections, risk predictions, etc. The desire for many as seen, is to address challenges with novel ML techniques from different perspectives. Differences in feature selection and optimization technique tools to estimate variable importance and to improve on prediction performance is also indicated with varying outcomes. Model performance evaluation is also indicated in almost all literature reviewed. However, there is a strong desire with few exceptions among majority of the reviewed literature to estimate best model performance significantly on prediction accuracy score irrespective of the problem domain. We also note the recorded higher value of balanced accuracy score by [56] achieved using up-sampling optimization technique from [54][55]. Within the healthcare system, the occurrence of dataset class imbalance for which the minority class maybe of outmost importance. As an example; in the prediction of patient treatment default; the number of non-defaulters may far exceed the number of defaulters by 100s of 1000s or in significant ratios such as 1:100000 but the ultimate key objective is to correctly identify those defaulters which maybe over-looked by most predictive algorithms. This is a major challenge in predictive modeling accuracy estimations. Therefore predictive modeling must take into account the skewedness in dataset classes so as to estimate reasonable prediction accuracies for a proper determination of best predictive technique.

In the light of the challenges identified, we propose a novel evaluation approach for predictive modeling use (shown in figure7 below) within healthcare systems context called PMEA (Proposed Model Evaluation Approach) which can be generalized in similar contexts. PMEA, addresses challenges for the use of prediction accuracy with balanced accuracy score derived from two most important evaluation metrics (True positive rates and True negative rates: TPR, TNR) to estimate more accurately best model performance in context. Graphical description of the processes that leads to the estimation of balanced accuracy score is shown in figure1 above.

**Figure7.**
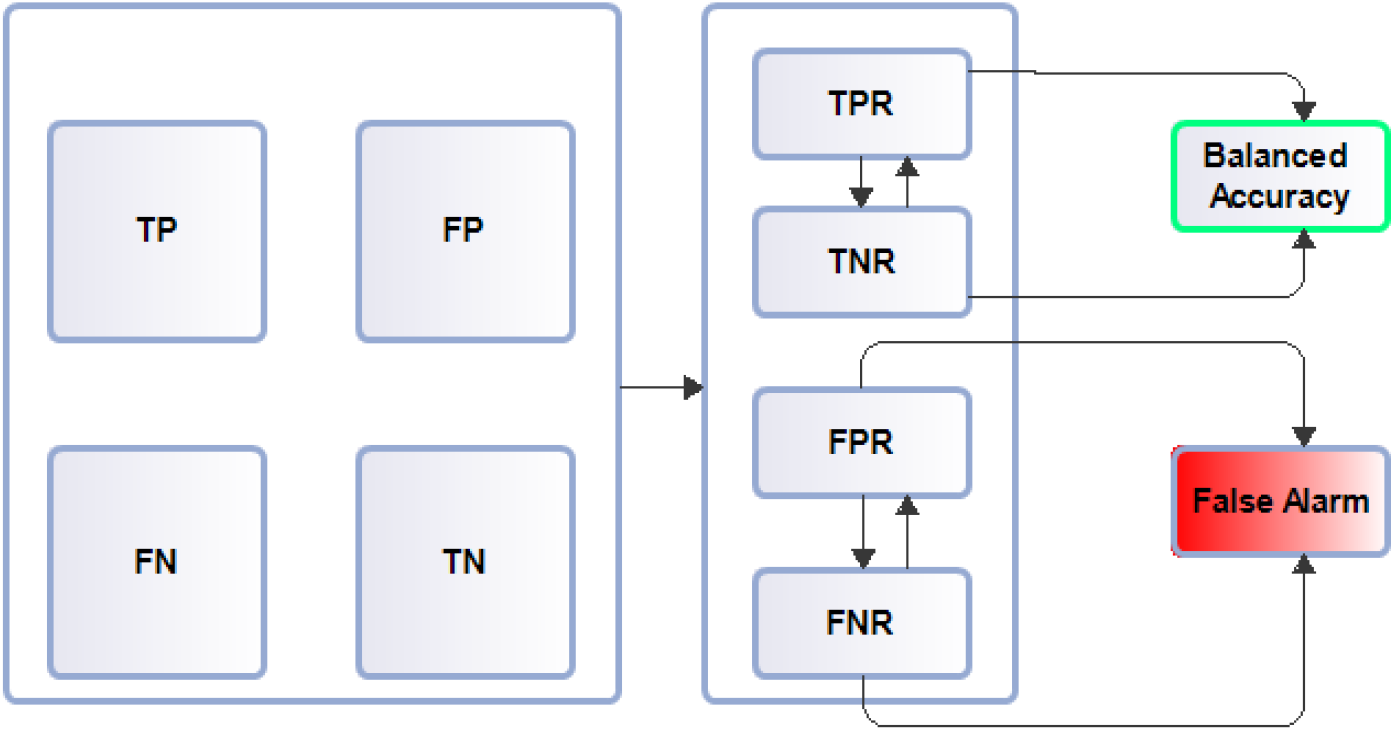
Evaluation process model.

### Proposed Evaluation Model

## Discussion

In this study, a review of related literature on the use of predictive modeling in real-world applications such as healthcare for either prediction of a certain disease or diagnosis of a disease and its related outcome. Approaches to estimate prediction outcomes have also been examined in the identified literature. Both strengths and weaknesses identified have been described. Challenges with approaches have been mentioned. This review is not the final determination of all the challenges in ML applications as, ML use is diverse and expanding so new challenges and opportunities will continue to emerge. While it may be fair to use prediction accuracy to justify model performance, its contextual application maybe understood than a generalization of prediction accuracy score use as the final evaluation metric to determine best model performance. Model performance in healthcare systems have a unique role to play. This is because lives are at stake. Assessing predictive performance based on probability score of false positives or false negatives (false alarms) within the healthcare system will be more beneficial to estimate best model performance than a focus on prediction accuracy score. And a prediction accuracy estimate based on the number of true positive and true negative rates will be a fair justification for estimating best model in performance within healthcare systems.

## Conclusion

We have examined literature, identified individual approaches to solving issues including context and examination of individual approaches. We have proposed an approach to deal with an identified challenge in context. This, we believe is not exhaustive, other evaluation assessments for its applicability in context will be examined in future research studies.

## Data Availability

Applicable upon request

## Acknowledgements

We acknowledge the support and cooperation of management and staff of Kwahu Government Hospital.

## Author contributions

**Conceptualization:** Owusu-Adjei Michael.

**Formal analysis:** Twum Frimpong.

**Methodology:** Twum Frimpong.

**Project administration:** Gaddafi Abdul-Salaam.

**Supervision:** James Ben Hayfron-Acquah.

**Writing – original draft:** Owusu-Adjei Michael.

**Writing – review & editing:** Twum Frimpong.

